# Associations between Neonicotinoid, Pyrethroid, and Organophosphate Insecticide Metabolites and Neurobehavioral Performance in Ecuadorian Adolescents

**DOI:** 10.1101/2024.10.10.24315201

**Authors:** Jessica Yen, Kun Yang, Xin M. Tu, Georgia Kayser, Ana Skomal, Sheila Gahagan, Jose Suarez-Torres, Suzi Hong, Raeanne C. Moore, Jose R. Suarez-Lopez

**Affiliations:** School of Medicine, University of California, San Diego, La Jolla, California, CA, 92093, USA; Herbert Wertheim School of Public Health and Human Longevity Science, University of California, San Diego, La Jolla, CA, 92093, USA; Department of Medicine, University of California, San Francisco, San Francisco, CA, 94143, USA; Department of Pediatrics, University of California, San Diego, La Jolla, CA, 92093, USA; Fundación Cimas del Ecuador, Quito, Ecuador; Department of Psychiatry, University of California, San Diego, La Jolla, CA, 92093, USA

**Author notes:** Corresponding Author: José Ricardo Suárez-López, MD, PhD, MPH, Herbert Wertheim School of Public Health and Human Longevity Science, University of California, San Diego, 9500 Gilman Drive #0725, La Jolla, California 92093-0725, USA.

**Keywords:** Insecticides, neurocognitive, pesticides, agriculture, floriculture, Rural Latin America

## Abstract

**Background:** Organophosphate and pyrethroid insecticides can affect children’s neurodevelopment and increase inflammation. Limited evidence exists among adolescents and on whether inflammation may mediate pesticide–neurobehavior associations. We examined the associations between insecticide metabolite concentrations and neurobehavior among adolescents in Ecuadorian agricultural communities.

**Methods:** We included 520 participants aged 11-17 years. We measured urinary insecticide metabolites (mass spectrometry) and neurobehavior (NEPSY-II). Associations were adjusted for socio-demographic and anthropometric characteristics. The associations of insecticide mixtures with neurobehavior were evaluated using PLS regression, and mediation by inflammatory biomarkers (TNF-α, IL-6, CRP, SAA, sICAM-1, sVCAM-1 and sCD-14) was conducted.

**Results:** Among organophosphates, para-nitrophenol (PNP) and 3,5,6-Trichloro-2-pyridinol (TCPy) were inversely associated with Social Perception (score difference per 50% increase [β_50%_] = -0.26 [95%CI: - 1.07, -0.20] and -0.10 [-0.22, 0.01], respectively). PNP and TCPy also had significant inverse associations with Attention/Inhibitory Control at concentrations >60^th^ percentile (β_50%_= -0.26 [95%CI: -0.51, -0.01] and β_50%_= -0.22 [95%CI: -0.43, -0.00], respectively). The pyrethroid, 3-phenoxybenzoic acid (3-PBA), was inversely associated with Language (β_50%_ = -0.13 [95%CI: -0.19, -0.01]) and had a negative quadratic association with Attention/Inhibitory Control. The neonicotinoid 5-Hydroxy imidacloprid (OHIM) was positively associated with Memory/Learning (β_50%_ = 0.20 [95%CI: 0.04, 0.37]). Mixtures of all insecticides were significantly negatively related to all domains, except for Memory/Learning, which was positively associated. No mediation by inflammatory markers on these associations was observed.

**Conclusions:** Concurrent organophosphate, pyrethroid, and the mixtures of all metabolites were associated with lower performance in all domains except for Memory/Learning. Neonicotinoids were positively associated with Memory/Learning and Social Perception scores.

## Introduction

With the global mass production of agriculture, insecticide use is becoming increasingly prevalent and widespread (Atwood and Paisley-Jones, 2017). There is substantial evidence that organophosphate insecticides can affect the neurobehavioral development of children. Organophosphates inhibit the acetylcholinesterase (AChE) enzyme, which results in acetylcholine accumulation in the body and can subsequently result in physical and neurological harm (Roberts and Karr, 2012). Prenatal exposure has been linked to impaired mental development (Eskenazi et al., 2008, 2007; Marks et al., 2010; Mostafalou and Abdollahi, 2017; Rauh et al., 2006; Vrijheid et al., 2016), reduced IQ (Gunier et al., 2017; Vrijheid et al., 2016), reduced verbal comprehension (Gunier et al., 2017), and increased rates of attention deficit hyperactivity disorder (ADHD) (Dalsager et al., 2019; Fortenberry et al., 2014) and autism spectrum disorder (ASD) (Mostafalou and Abdollahi, 2017; Sagiv et al., 2018; Shelton et al., 2014) in early childhood. Studies regarding postnatal organophosphate exposures have been less consistent (Li et al., 2012; Vrijheid et al., 2016), but they suggest an association between childhood organophosphate exposure (measured by urinary insecticide metabolite levels and/or reduced AChE activity) and impaired mental development, verbal function, memory, attention, and inhibitory control as well as the development of ADHD and ASD (Butler-Dawson et al., 2016; Eskenazi et al., 2007; Ismail et al., 2017b; Jurewicz et al., 2013; Kofman et al., 2006a; Mostafalou and Abdollahi, 2017; Rohlman et al., 2016; Ruckart et al., 2004; Suarez-Lopez et al., 2013).

Pyrethroid insecticides are considered less harmful and more varied in their biological effects than organophosphates are (Roberts and Karr, 2012). However, pyrethroids are known to inhibit nerve cell function (Roberts and Karr, 2012; Saillenfait et al., 2015) and alter brain dopaminergic activity (Carloni et al., 2012; Elwan et al., 2006; Nasuti et al., 2013, 2007; Richardson et al., 2015). Prenatal exposure to environmental pyrethroids has been associated with neurodevelopmental deficits (Horton et al., 2011; Saillenfait et al., 2015; van Wendel de Joode et al., 2016; Xue et al., 2013), lower IQ (Gunier et al., 2017), ASD (Mostafalou and Abdollahi, 2017), and ADHD symptoms (Dalsager et al., 2019) in children from urban and agricultural settings. Limited evidence suggests that postnatal pyrethroid exposure can also affect neurodevelopment (e.g., memory, verbal comprehension, social behaviors) (Viel et al., 2017; Wang et al., 2016) and increase the rates of behavior problems (Oulhote and Bouchard, 2013) and ADHD in adolescents, especially those living near agricultural crops (van Wendel de Joode et al., 2016).

Neonicotinoids have become the most commonly used class of insecticides (Jeschke and Nauen, 2008; Simon-Delso et al., 2015). However, few studies have evaluated the neurobehavioral effects in humans (Cimino et al., 2017; Han et al., 2018; Ongono et al., 2020). Neonicotinoids are nicotinic acetylcholine receptor (nAChR) agonists, a mechanism that provides insecticide activity but can also cause neurotoxicity in mammals (Anadón et al., 2020; Han et al., 2018). In mammalian studies, neonicotinoid exposure in neonatal rats resulted in neurobehavioral and sensorimotor deficits by adolescence (Anadón et al., 2020). Acute and subacute neonicotinoid exposure in adult rats is associated with neurologic and motor symptoms (Anadón et al., 2020). In humans, prenatal insecticide exposure, assessed through geospatial analysis of prenatal proximity to agricultural insecticide sites (Gunier et al., 2017) and interviews regarding prenatal household neonicotinoid use (Keil et al., 2014), has been associated with reduced IQ (Gunier et al., 2017) and ASD in childhood (Keil et al., 2014). Urinary neonicotinoid levels have been linked to neurologic symptoms, including memory loss, finger tremor, and headache, in children and adults (Cimino et al., 2017; Han et al., 2018).

A limited number of human studies suggest that pesticide exposure may result in inflammatory alterations. For example, higher concentrations of urinary organophosphate and pyrethroid metabolites are associated with higher levels of various interleukins (ILs: 4, 5, 6, 8, 10, 13 and 17) (Mwanga et al., 2016). In our prior work in adolescents in agricultural communities in Ecuador, we reported that a mixture of insecticides, herbicides and insect repellents were positively associated with various inflammatory biomarkers, including IL-6, serum amyloid A (SAA), tumor necrosis factor (TNF-llJ), soluble intercellular adhesion molecule-1 (sICAM-1) and soluble vascular cell adhesion molecule-1 (sVCAM-1) (*Mohamed N. Hussari, article currently under review*). In this same population, we also recently reported that a composite of inflammation markers, including C-reactive protein (CRP), IL-6, TNF-llJ, sICAM-1, sVCAM-1, SAA, and soluble CD-14 (sCD-14), was negatively associated with neurobehavioral performance in the domains of language, visuospatial processing and social perception (*Beemnet Amdemicael, et al. article currently under review*). Inflammation could be a mechanism by which pesticides impact neurobehavioral performance, making it an important area for further study.

Insecticide exposure is a health concern for agricultural workers; however, family members and others are also at risk for contamination through environmental, dietary, and take-home pathways (Butler-Dawson et al., 2016; Crane et al., 2013; Hung et al., 2018; Morgan, 2012; Panuwet et al., 2009; Saillenfait et al., 2015; Suarez-Lopez et al., 2012; Zhang et al., 2018). Insecticide metabolites have been detected in the urine of agricultural workers (Abdel Rasoul et al., 2008; Ismail et al., 2017b; Tao et al., 2019b) as well as in the general population (Han et al., 2018; Hill et al., 1995; Panuwet et al., 2009; Wielgomas and Piskunowicz, 2013) and in most children living in urban and agricultural communities (Morgan, 2012).

Compared with those living further from crops, exposure levels are particularly high among children living closest to farms (Friedman et al., 2020; Tao et al., 2019a; Wielgomas and Piskunowicz, 2013). Gender may also influence the neurobehavioral effects of insecticide exposure. In boys but not girls, exposure to organophosphates and pyrethroids has been associated with deficits in attention and inhibitory control (Lee et al., 2020; Marks et al., 2010; Richardson et al., 2015; Suarez-Lopez et al., 2013; Wagner-Schuman et al., 2015; Yu et al., 2016) and memory problems (Horton et al., 2012; Suarez-Lopez et al., 2013). Neonicotinoids have demonstrated differing toxicity profiles between male and female mammals (Anadón et al., 2020). Additionally, boys are more susceptible than girls to neurobehavioral deficits related to periods of peak pesticide exposure(Suarez-Lopez et al., 2017a).

Adolescence is a period characterized by extensive physiological and brain maturation changes in which many mental health disorders are initiated. Given the limited number of studies in adolescents that have assessed agrochemical exposure and neurobehavior, the present study aimed to examine the relationships between urinary insecticide metabolites (i.e., organophosphates, pyrethroids, and neonicotinoids) and performance in various neurobehavioral domains among adolescents aged 12--17 years living in agricultural settings in Ecuador. On the basis of the existing data, we hypothesized that urinary organophosphate and pyrethroid concentrations would be inversely associated with the domains of Attention & Inhibitory control, Language, Memory & Learning, and Social Perception. Though the evidence for neonicotinoids is less clear, we hypothesized that neonicotinoids would have negative associations with Memory & Learning and Social Perception. A second aim was to identify whether these relationships differed by gender. We hypothesized that the associations between insecticide metabolites and neurobehavioral deficits would be stronger for boys compared to girls, particularly in the domains of Attention & Inhibitory Control and Memory & Learning. Finally, we investigated the potential mediating role of inflammation in the effects of insecticides on neurobehavior as a preliminary step toward understanding the underlying mechanisms involved.

## Methods

Data for this study were obtained from the longitudinal Secondary Exposure to Pesticides among Children Adolescents and Adults study (ESPINA – Estudio de la Exposición Secundaria a Plaguicidas en Niños, Adolescentes y Adultos). The overall aims of ESPINA are to examine the association of pesticide exposure with the development of children living in the agricultural county of Pedro Moncayo, Pichincha, Ecuador. Pedro Moncayo County is located in the Ecuadorian Andes and hosts a large floricultural industry in which more than 20 different insecticides, including organophosphates, carbamates, neonicotinoids, and pyrethroids, are used (Suarez-Lopez et al., 2018). Children living in this agricultural community have shown biological signs of exposures to organophosphates, especially those living closest to flower plantations (Suarez-Lopez et al., 2018; Suárez-López et al., 2020) or cohabiting with flower workers (Suarez-Lopez et al., 2012). The ESPINA study began in 2008, at which time 313 children aged 4-9 years were enrolled. Seventy-three percent of these children were recruited from the 2004 Survey of Access and Demand of Health Services in Pedro Moncayo County, which collected data on 71% of the county’s population. This survey was carried out in Spanish by Fundación Cimas del Ecuador in collaboration with the Local Rural Governments of Pedro Moncayo and community residents. The other 27% of participants were recruited through community announcements and word-of-mouth. Children were included in the study if they (1) lived with flower workers or cohabited with a flower plantation worker for at least one year or (2) lived without an agricultural worker, never cohabited with an agricultural worker, never lived in a house that stored agricultural pesticides, and had no prior direct contact with pesticides. Families received financial compensation for their participation in the study.

In 2016, we reexamined 238 participants who were enrolled in 2008 plus an additional 316 new participants, for a total of 554 adolescents aged 11--17 years. Among the 554 participants, 535 were examined between July and October 2016, and 330 were examined in April. There were 311 participants who were examined in both the April and July–October exams. New participants were recruited via the System of Local and Community Information (SILC), a geocoded database developed by Fundación Cimas del Ecuador containing information from the 2016 Pedro Moncayo County Community Survey (formerly the Survey of Access and Demand of Health Services in Pedro Moncayo County). Participant recruitment in 2008 and 2016 is described in more detail elsewhere (Suarez-Lopez et al., 2012, 2019a). The current analysis included 520 out of 535 (97%) adolescents examined in the July–October exam in 2016 who had information on all covariates and neurobehavioral outcomes of interest. Among the study participants in 2016, none worked in agriculture, and 67% lived with an agricultural worker (Chronister et al., 2023). Multiple children per household were allowed to participate.

### Ethical approval and informed consent

Informed consent and parental permission for child participation were obtained from at least one parent of each child, and written consent was obtained from all the children over 7 years of age. This study was approved by the institutional review boards at the University of California San Diego, Universidad San Francisco de Quito, and was approved by the Ministry of Public Health of Ecuador.

### Household survey

Parents and other adult residents were surveyed at home about socioeconomic status, demographic characteristics, health, and pesticide exposure of household members.

### Examination

In 2016, adolescents were examined on two occasions. Examinations took place across seven schools in Pedro Moncayo County. In April, 330 children completed an abbreviated examination which included some subtests of the NEPSY-II assessment (see below). Between July and October, during the summer school closure or on weekends, 535 children participated in a more comprehensive examination. which included both the NEPSY-II test and urine pesticide biomarker measures. The examiners were unaware of the participants’ pesticide exposure status. Height was measured to the nearest 1 mm via a height board following recommended procedures (Organization, 2008), and weight was measured via a digital scale (Tanita model 0108 MC; Corporation of America, Arlington Heights, IL, USA). We calculated height-for-age z score (z-height-for-age) and body mass index-for-age z score (z-BMI-for-age) using the World Health Organization (WHO) growth standards (WHO Multicentre Growth Reference Study Group, 2006). Sexual maturation rating (SMR) was assessed based on self-reported breast size and pubic hair growth/distribution for girls, and pubic hair growth/distribution for boys, using modified Tanner drawings used as a reference (Emmanuel et al., 2020; Kormorniczak, 2009; Rasmussen et al., 2015).

### Neurobehavioral testing

Assessments were administered using the NEPSY-II test (NCS Pearson, San Antonio, TX), a standardized and validated neurodevelopment test battery for children aged 3–16 years (Kemp and Korkman, 2010; Korkman et al., 2007a). Participants were examined in a quiet room by a trained examiner. Only the participant and an examiner were allowed in the room. During the July– October 2016 examination, the adolescents were tested in nine age-appropriate subtests, corresponding to five domains: 1) Attention & Inhibitory Control (also known as Attention and Executive Functioning, subtests: auditory attention and response set, inhibition); 2) Language (subtests: comprehension of instructions, speeded naming); 3) Memory & Learning (subtests: memory-for-faces immediate, memory-for-faces delayed); 4) Visuospatial Processing (subtests: design copying, geometric puzzles); and 5) Social Perception (subtest: affect recognition). Descriptions of each subtest can be found elsewhere (Korkman et al., 2007a; Suarez-Lopez et al., 2013). Two subtests (auditory attention and response set and comprehension of instructions) required translation into Spanish. Translations used terminology appropriate for the local population and were approved by the NCS Pearson. Translation of the NEPSY test has been found to be relatively unaffected by language and culture (Garratt and Kelly, 2008; Kofman et al., 2006b; Mulenga et al., 2001). NEPSY-II scaled scores for each subtest can range from 1--19 and have a mean of 10 (SD=3), which is based on a US normative sample (Brooks et al., 2010; Korkman, M., Kirk, U., & Kemp, 2007). Most adolescents completed the general assessment battery within 50--80 minutes. Attention & Inhibitory Control were also assessed in April 2016 among 303 participants. For these participants, the models were further adjusted for test-retest learning effects in this domain (see statistical analysis).

### Biospecimen collection, storage and transportation

Urinary concentrations of creatinine and insecticide biomarkers were measured in samples collected upon awakening from July October 2016. The participants brought the urine samples to the examination site in the morning, where they were aliquoted and frozen at -20°C. At the end of each day, the samples were transported to Quito for storage at -70°C. The samples were then transported overnight to the UCSD at -20°C via a courier and stored at -80°C at the UCSD. The samples were then shipped overnight at -20 ^°^C from the UCSD to the National Center for Environmental Health, Division of Laboratory Sciences of the CDC (Atlanta, GA) for the quantification of organophosphate, pyrethroid and neonicotinoid metabolites. Quality control/quality assurance protocols were followed to ensure the data accuracy and reliability of the analytical measurements. All the study samples were re-extracted if quality control failed the statistical evaluation (Baker et al., 2019).

### Organophosphate and pyrethroid metabolite quantification

Urinary concentrations of organophosphates (malathion dicarboxylic acid (MDA), 3,5,6-trichloro-2-pyridinol (TCPy), 2-isopropyl-4-methyl-6-hydroxypyrimidine (OXY2), and *para*-nitrophenol (PNP)) as well as pyrethroids (4-fluoro-3-phenoxybenzoic acid (4FP), 3-phenoxybenzoic acid (3-PBA), and trans-3-(2,2-dichlorovinyl)-2,2-dimethylcyclopropane carboxylic acid (trans-DCCA)) were quantified. Targeted metabolites were extracted from 1.5 mL of urine via semiautomated solid-phase extraction, removed from each other and other urinary biomolecules via reversed-phase high-performance liquid chromatography, and identified via isotope dilution quantitation via tandem mass spectrometry (Davis et al., 2013). Quality control measures were followed for each step for an accurate, precise, reproducible, and efficient method. The limits of detection (LODs) were 0.5 µg/L (MDA), 0.1 µg/L (TCPy), 0.1 µg/L (OXY2), 0.1 µg/L (PNP), 0.1 µg/L (4FP), 0.1 µg/L (3-PBA), and 0.6 µg/L (trans-DCCA).

### Neonicotinoid metabolite quantification

The neonicotinoids Acetamiprid-N-desmethyl (AND), 5-Hydroxy imidacloprid (OHIM), Acetamiprid (ACET), Clothianidin (CLOT), Imidacloprid (IMI), and Thiacloprid (THIA) were quantified from 0.5 mL of urine via enzymatic hydrolysis of urinary conjugates of target biomarkers and online solid-state extraction, separated by reversed-phase high-performance liquid chromatography, and detection by isotope dilution-electrospray ionization tandem mass spectrometry. These methods are explained in more detail elsewhere (Baker et al., 2019). Strict quality control/quality assurance protocols were followed to ensure data accuracy and reliability for the analytical measurements. If the quality control samples failed the statistical evaluation, all the study samples were re-extracted (Caudill et al., 2008). The LODs were 0.2 μg/L (AND), 0.4 μg/L (OHIM), 0.3 µg/L (ACET), 0.1 µg/L (CLOT), 0.4 µg/L (IMI), and 0.03 µg/L (THIA).

### Creatinine

Urinary creatinine was quantified using HPLC-MS/MS with ESI at the Laboratory for Exposure Assessment and Development in Environmental Research at Emory University (Atlanta, GA). A 10 μL aliquot of urine was diluted prior to analysis (Kwon et al., 2012). No further sample preparation was performed prior to analysis. The LOD was 5 mg/dL, with an RSD of 7%.

### Inflammatory markers

Venipuncture of the median cubital or cephalic veins (arm) was conducted by a phlebotomist following standard guidelines (Clinical and Laboratory Standards Institute, 2007). The participants were not asked to fast prior to venipuncture. Blood samples were analyzed for inflammation marker quantification at the UCSD Integrative Health Mind-Body Biomarker Core. Multiplex assay kits were used to measure the plasma levels of TNF-α, IL-6, CRP, SAA, sICAM-1, and sVCAM)-1 via a 2400 SECTOR Imager reader (Meso Scale Discovery, Rockville, Maryland). The sCD-14 levels were measured using quantikine ELISA kits from R&D Systems (Minneapolis, MN). The intra- and inter assay coefficients of variation of those markers were less than 10%. The inflammation summary score was calculated by averaging the z scores of seven log-transformed inflammatory markers.

### Geospatial

The distance from the participants’ homes to the nearest flower plantation was calculated via ArcGIS 9.3 (Esri, Redlands, CA) from geographical coordinates obtained from portable global positioning system receivers and satellite imagery. Detailed data collection information has been described elsewhere (Suarez-Lopez et al., 2012).

## Statistical analysis

### Urinary metabolites

To calculate the percentage of participants in which each metabolite was detectable, we used a denominator that excluded participants with interfering substances that precluded the measurement of the metabolites in the urine. *Imputation of values below the limit of detection (LOD).* For metabolite concentrations below the LOD, we used two different imputation methods. Method 1 assigned a fixed value (LOD/square root of 2) to metabolite concentrations below the LOD. Method 1 was used for metabolites detected in fewer than 25% of the participants and only for the calculation of pesticide summary scores. Method 2 was used for metabolites that were detectable in 25% or more of the participants. The log linear regression method was used for the prediction of the imputed values. The imputation models were fitted by backward selection with a threshold of a p-value less than 0.1 based on the observed values of the urinary concentration as the response. There is a large set of covariates used for the initial model, including the urinary concentrations of TCPy and PNP, gender, age, race, creatinine, hemoglobin, z-height-for-age, z-BMI-for-age, Tanner score, salary, area of flower crops within 150 meters of the participant’s home, child cohabitation status with agricultural workers, residential distance to nearest flower plantation, and acetylcholinesterase activity. The imputation model for MDA included gender, race, creatinine, hemoglobin, cohabitation with an agricultural worker, and the TCPy concentration. The imputation model for 3-PBA included age, race, creatinine, z-BMI-for-age, and TCPy concentration. The imputation model for AND included gender, race, z-BMI-for-age, cohabitation status, and acetylcholinesterase activity. The imputation model for OHIM included salary, residential distance to nearest flower plantation, and TCPy concentrations. For an analysis that includes imputed data, we treated the observed values and imputed values as two variables in one model so that there was no need for rescaling the imputed values. The missing values for each side were set as 0 or 1 if log-transformation was needed. All insecticide metabolite concentrations were natural log-transformed due to excessive skewness. We also created an indicator variable with 1 for the imputed values and 0 for the observed values so that the two groups could have different intercepts. *Summary scores.* Because populations tend to be exposed to pesticide combinations as opposed to individual pesticides in isolation (Porta et al., 2008) and our study participants exhibited varying insecticide exposure levels, we created summary scores reflecting overall insecticide exposure (organophosphates, pyrethroids, and neonicotinoids combined) as well as categorical exposure to organophosphates (MDA, TCPy, PNP, OXY2), pyrethroids (3-PBA, trans-DCCA), and neonicotinoids (AND, OHIM, ACET, CLOT, IMI) for each participant. Categorical exposure is an estimate of exposure to an individual group of insecticides (organophosphates, pyrethroids, or neonicotinoids) and accounts for exposure to each of the insecticide subtypes within the corresponding category. In our calculation of categorical exposure, in addition to insecticides detected in 25% or more of the participants, we included insecticides (OXY2, trans-DCCA, ACET, CLOT, IMI) detected in fewer than 25% of the participants, who were excluded from our overall analysis of the associations between insecticide concentration and neurobehavioral performance. For the insecticides detected in fewer than 25% of the participants, we included the observed values and fixed imputed values for concentrations below the LOD. For the insecticides detected in 25% or more of the participants, we included the observed values and the backward selected imputed values for concentrations below the LOD. As 4FP (pyrethroid) and THIA (neonicotinoid) were both detectable in 0% of the participants, they were excluded from the summary scores. We also excluded metabolite concentrations that were unobtainable due to an interfering substance in the code. Summary scores were calculated by first adding 1 to the observed and imputed participant metabolite concentrations and then log transforming the new metabolite values to create a more normal distribution. We added 1 to the original concentrations to ensure that all log-transformed values were positive. We divided these values (s = log[metabolite concentration + 1]) by the group’s standard deviation of the corresponding log-transformed metabolite concentrations. To obtain a summary score, we took the average of the normalized results: 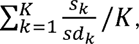 where k refers to the *k*th metabolite, K refers to the total number of metabolites in the metabolite class and sd denotes the standard deviation of s.

### Neurobehavior scores

Subtest-scaled scores, which are age-adjusted values based on a national normative sample of children/adolescents in the United States (Korkman et al., 2007b), were calculated via the NEPSY-II scoring assistant software (NCS Pearson Inc., San Antonio, TX). Higher scores reflect better performance. The scaled scores range from 1 to 19 (mean=10, SD=3). A higher score on the NEPSY-II indicates better neurobehavioral performance, where scores 13 to 19 is “above expected”, 8 to 12 is “at expected”, 6 to 7 is “borderline” and less than 6 is “below expected”.(Korkman et al., 2007b) Most subtest scaled scores consisted of the subtest’s primary scaled score. For subtests that included time and error components (inhibition, speeded naming, visuomotor precision) and correct and error components (AARS), we used the combined scaled scores (scores that combined both components) as primary scaled scores. In subtests that included >1 subtest component and provided >1 primary scaled score (AARS, inhibition), each component primary score was analyzed separately (for AARS: AARS– auditory attention and AARS–response set; for inhibition: inhibition-naming [IN-naming] trial, inhibition-inhibition [IN-inhibition] trial, and inhibition-switching [IN-switching] trial). We excluded IN-naming trials from the analyses because this component assesses only the ability to name figures, which does not in itself reflect inhibitory control. We calculated domain scores by averaging the primary scaled scores from all subtests within each domain. For subtests with more than one primary scaled score, we first calculate the average of the primary scaled scores for the subtest and then incorporate that value in the domain calculation. Since the Social Perception domain consists of only one subtest (affect recognition), the domain score is equal to the subtest score. Additional details of subtest scoring have been published elsewhere (Korkman, M., Kirk, U., & Kemp, 2007; Suarez-Lopez et al., 2013). *Adjusting for test-retest learning effects.* There were 303 adolescents who completed the Attention & Inhibitory Control domain of the NEPSY-II twice in 2016 (in April and summer [July-October]). To account for learning or practice effects in repeat test takers, we categorized participants into two groups. The Retest group consisted of children who were examined in both April and Summer exams, and the non-retest group consisted of children who were only examined in the summer. When comparing the mean scores of the summer assessments, we found that the retest group had higher scores than did the non-retest group (8.92 compared with 7.60, difference p value <0.0001), confirming the presence of a learning effect in those taking the NEPSY-II for the second time. Since practice effect only applied to the Attention & Inhibitory Control domain of the NEPSY-II, we controlled for practice effect in the analyses of Attention & Inhibitory Control domain subtests only. To do this, we used an indicator variable to differentiate participants who had taken the assessment before from those who were taking the assessment for the first time. We then adjusted for this indicator variable in our models.

### Generalized estimating equations (GEEs)

We analyzed the associations of individual insecticide metabolites and insecticide summary scores (overall, organophosphates, pyrethroids, neonicotinoids) with neurodevelopment scores via GEE models (Tang et al., 2012). We evaluated curvilinear associations via quadratic terms (β + β^2^). The models were adjusted for age, gender, race, creatinine, z-height-for-age, z-BMI-for-age, hemoglobin concentration, Tanner score, salary, and parental education. The beta coefficients calculated by the model were multiplied by log(1.50). With this adjustment, the coefficients in Table 3 can be interpreted as the amount of change in the dependent variable (neurobehavioral domain score) corresponding to a 50% increase in the independent variable (metabolite concentration). We used a multiple imputation method for the estimate of the coefficients and p values if the model included the imputed values. The procedure of generating imputed values by introducing a random error with a standard normal distribution and fitting the GEE model with imputed data was repeated 1,000 times. The mean and standard deviation of each coefficient can be estimated with the 1,000 fitted models. We assessed effect modification by gender on the pesticide-neurobehavior association by testing the statistical significance of a multiplicative term (gender*metabolite concentration). For associations that showed evidence of gender interaction at p<0.10, we stratified our analysis by gender.

Locally weighted polynomial regression (LOESS) curve graphs were created to visualize associations and to visually assess the presence of a threshold effect between insecticide metabolites and neurobehavioral performance. The presence of a threshold is defined as the presence of a significant association between the exposure and the outcome variables but only above a certain level of exposure. The LOESS graphs plotted the associations and 95% confidence intervals (smoothness factor: 0.9) via the fully adjusted least squares mean of each domain score for up to 300 ranks of observed insecticide metabolite concentrations and imputed concentrations. All analyses were performed via R statistical programming software. When the LOESS figures showed an indication of a possible threshold effect, we stratified the metabolite– neurobehavior associations by values above and below the visualized threshold.

### Partial least squares regression

We created composite variables from multiple pesticide metabolites, including TCPy, PNP, MDA, 3-PBA, AND and OHIM, to enhance the power of modeling their associations with neurobehavioral outcomes. For concentrations below the LOD, we included the imputed values generated from the multiple imputation models as well. For each imputed data point, the mean of the 1,000 repeats was calculated and then rescaled to a value between 0 and the LOD of the pesticide. The inflammation biomarker concentrations were standardized for PLS regression to eliminate scale differences between them. PLS generates composite variables that are linear combinations of pesticide metabolite concentrations, considering the relationship with each dependent variable separately. Multiple component variables are created, with the first composite having the maximum correlation with the dependent variable, followed by the second, and so on. PLS composite variables are effective for identifying a subset of explanatory variables (pesticide metabolites) in the linear model that explains the most variability in the response. We calculated the loadings of the composite variables for PLS component 1, which represent the differential contributions of each independent pesticide metabolite. A positive loading of a metabolite indicates that it aligns with the direction of the composite variable when interpreting the association of the biomarker with the dependent outcome in the regression analysis. To evaluate model fit, adjusted R-square values were calculated for each PLS component. A residualization procedure was used to adjust for covariates. This means that for each cognitive outcome, the outcome variable was represented by the residual obtained from a linear model that was fitted with the original outcome and the covariates.

### Mediation

As an exploratory analysis, we examined whether inflammatory biomarkers are mediators of pesticide–neurobehavior associations. To reduce the number of models, structural equation modeling (SEM) was applied to test whether the ‘inflammation summary score’, calculated as the mean of z scores of log-transformed levels of inflammatory markers, mediated the associations between the pesticide PLS component 1 and each neurobehavioral domain. We calculated direct, indirect, and total effects and expressed the indirect effect as a percentage to indicate the degree of mediation. Full mediation implies a direct effect of 0% and an indirect effect of 100%. These models were adjusted for the same covariates as the GEE models. We considered that mediation may be present if the indirect effect was statistically significant (p<0.05) or borderline significant (p<0.1). These analyses were performed using the R lavaan package.

## Results

### Participant characteristics

Among our participants, 51.0% were female, and 21.9% were indigenous (Table 1). The mean age was 14.5 years +/-1.8 years. Family income, mestizo ethnicity, and creatinine were positively associated with the overall insecticide summary score (p<0.05). Additional participant characteristics are listed in Table 1. The mean scores (SD) for the following neurodevelopmental domains were: Attention & Inhibitory Control: 8.37 (2.16), Language: 7.04 (2.05), Memory & Learning: 8.31 (2.29), Visuospatial Processing: 8.71 (2.26), Social Perception: 8.23 (2.34). Boys (mean score of 7.22) performed better than girls did (mean score of 6.86) in the Language domain (p_difference_= 0.05). However, performance in the other neurobehavioral domains did not significantly differ between boys and girls. The organophosphates TCPy and PNP were detectable in all participants. Interfering substances that precluded the measurement of PNP were present in the samples of five participants. The percentages of participants with measurable levels of metabolites (percent detectable) are presented in Table 2. We also present the number of participants who had interfering substances that precluded the measurement of these chemicals in the urine.

**Table 1.**
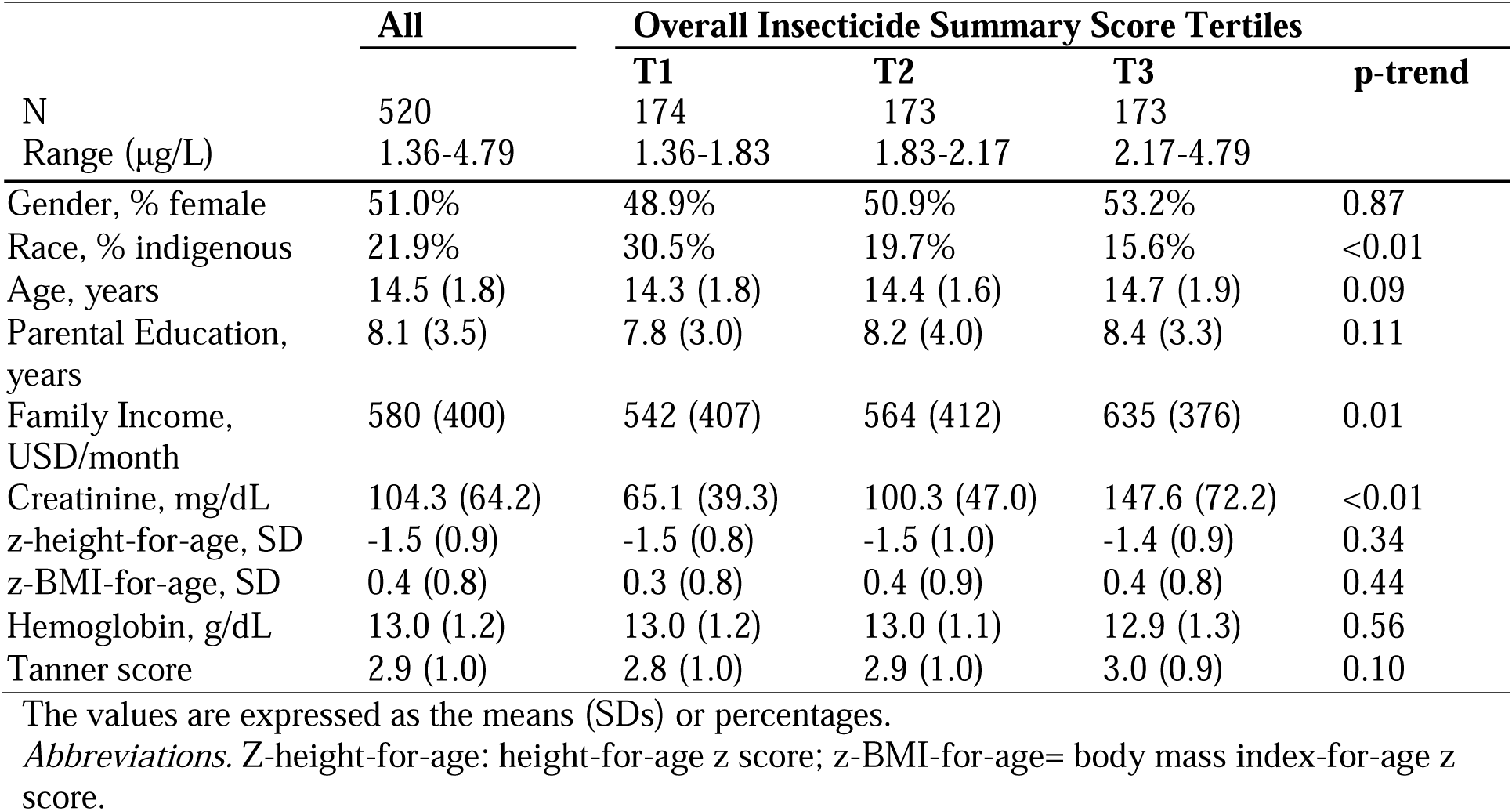
Participant characteristics across tertiles of overall insecticide summary score among participants examined from July–October 2016.

**Table 2.** Metabolite concentrations among participants examined in July-October 2016, N=520.

|  | **Percent Detectable, %** | **Interfering substances, N** | **Geometric Mean (95% CI), ug/L** |
| --- | --- | --- | --- |
| **Organophosphates** |  |  |  |
| TCPy | 100 | 0 | 2.69 (2.51, 2.88) |
| PNP | 100 | 5 | 0.52 (0.49, 0.54) |
| MDA | 37.7 | 0 | 0.50 (0.48, 0.53) |
| OXY2 | 23.7 | 3 | 0.10 (0.09, 0.11) |
| **Pyrethroids** |  |  |  |
| 3-PBA | 84.3 | 26 | 0.40 (0.35, 0.42) |
| Trans-DCCA | 15.9 | 12 | 0.53 (0.50, 0.55) |
| 4FP | 0.00 |  |  |
| **Neonicotinoids** |  |  |  |
| ACET | 1.1 | 1 | - |
| AND | 36.6 | 18 | 0.27 (0.24, 0.28) |
| CLOT | 2.8 | 0 | - |
| IMI | 7.6 | 10 | - |
| OHIM | 26.4 | 41 | 0.44 (0.38, 0.43) |
| THIA | 0.00 |  | - |

*Abbreviations.* Q1: 25<sup>th</sup> percentile, Q3: 75<sup>th</sup> percentile, TCPy: 3,5,6-trichloro-2-pyridinol, PNP: para-nitrophenol, MDA: malathion dicarboxylic acid, OXY2: 2-isopropyl-4-methyl-6-hydroxypyrimidine, 3-PBA: 3-phenoxybenzoic acid, trans-DCCA: *trans*-3-(2,2-Dichlorovinyl)-2,2-dimethylcyclopropane carboxylic acid, ACET: acetamiprid, AND: Acetamiprid-N-desmethyl, CLOT: clothianidin, IMI: imidacloprid, OHIM: 5-Hydroxy imidacloprid, 4FP: 4-fluoro-3-phenoxybenzoic acid

### Urinary Metabolite Concentrations and Neurobehavioral Development

Table 3 and Figures 1-4 show the relationships of insecticide summary scores and individual insecticide metabolites with neurobehavioral domain performance. The overall insecticide summary score was inversely associated with Language (score difference per 50% increase in metabolite concentration [β_50%_] = -0.33 [95% CI: -0.72, 0.05, p=0.09]) and Social Perception (β_50%_ = -0.50 [95% CI: -1.02, 0.022, =0.06]), with borderline non-significance. Among organophosphates, the organophosphate summary score was inversely associated with Social Perception (β_50%_ = -0.33 [95%CI: -0.60, -0.05, p=0.02]) and TCPy was inversely associated with Language (β_50%_ = -0.10 [95%CI: -0.19, -0.01, p=0.04]) and Social Perception (β_50%_ = -0.10 [95%CI: -0.22, 0.01, p=0.08]). TCPy also had a significant curvilinear association with Attention & Inhibitory Control (ß_quadratic_ = -0.21 [95%CI: -0.41, 0.00, p=0.05]) (Table 3). PNP also was inversely associated with Social Perception (β_50%_ = -0.26 [95%CI: -0.43, -0.08, p<0.01]), in addition to Attention & Inhibitory Control with borderline significance (β_50%_ = -0.12 [95%CI: -0.26, 0.02, p=0.09]). MDA had a significant curvilinear association with Memory & Learning (ß_quadratic_ = 0.72 [95%CI: 0.24, 1.20, p<0.01]) (Table 3).

**Figure 1.**
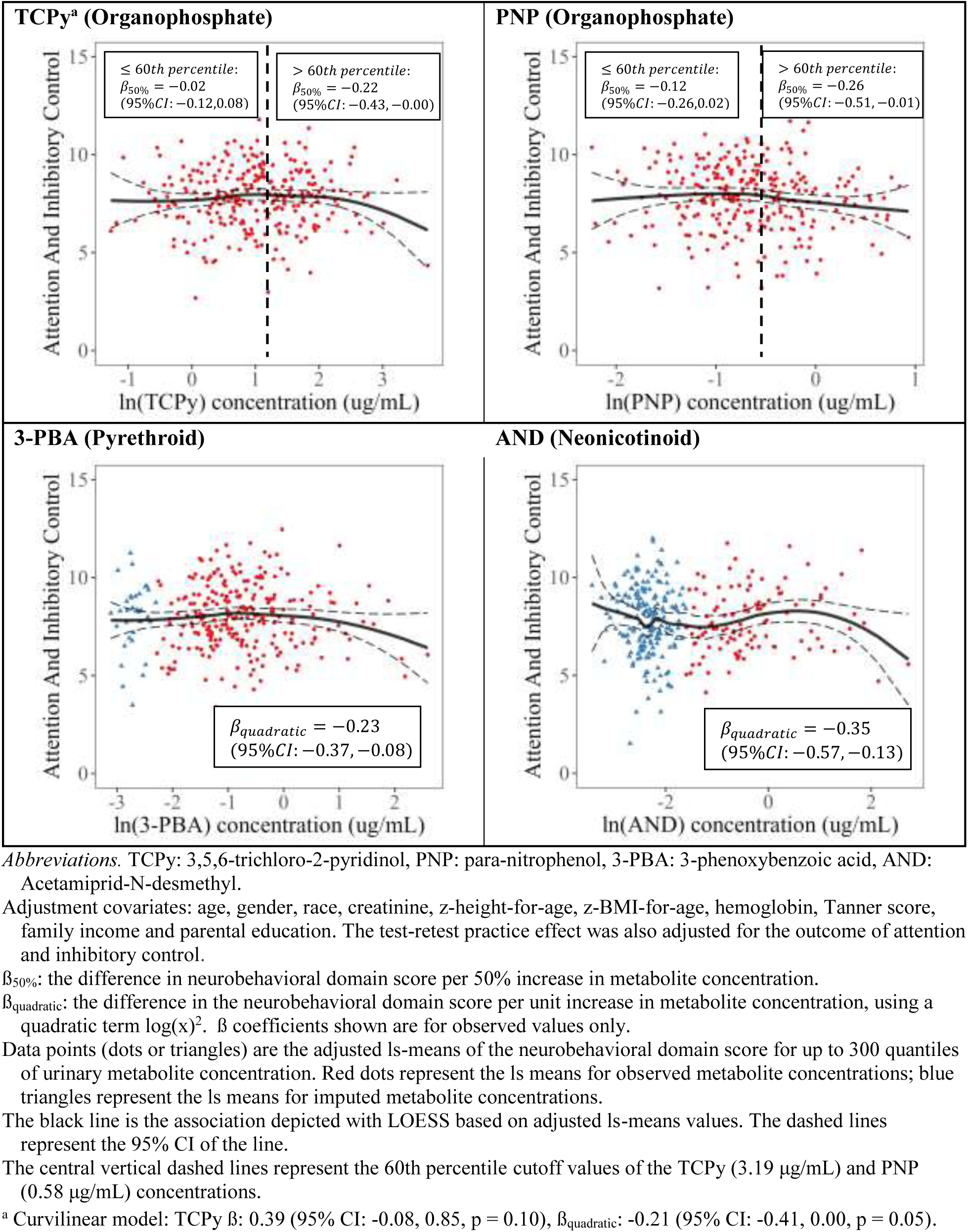
Attention and inhibitory control domain associations with log-transformed insecticide metabolites. Selected associations with p-values < 0.1.

**Figure 2.**
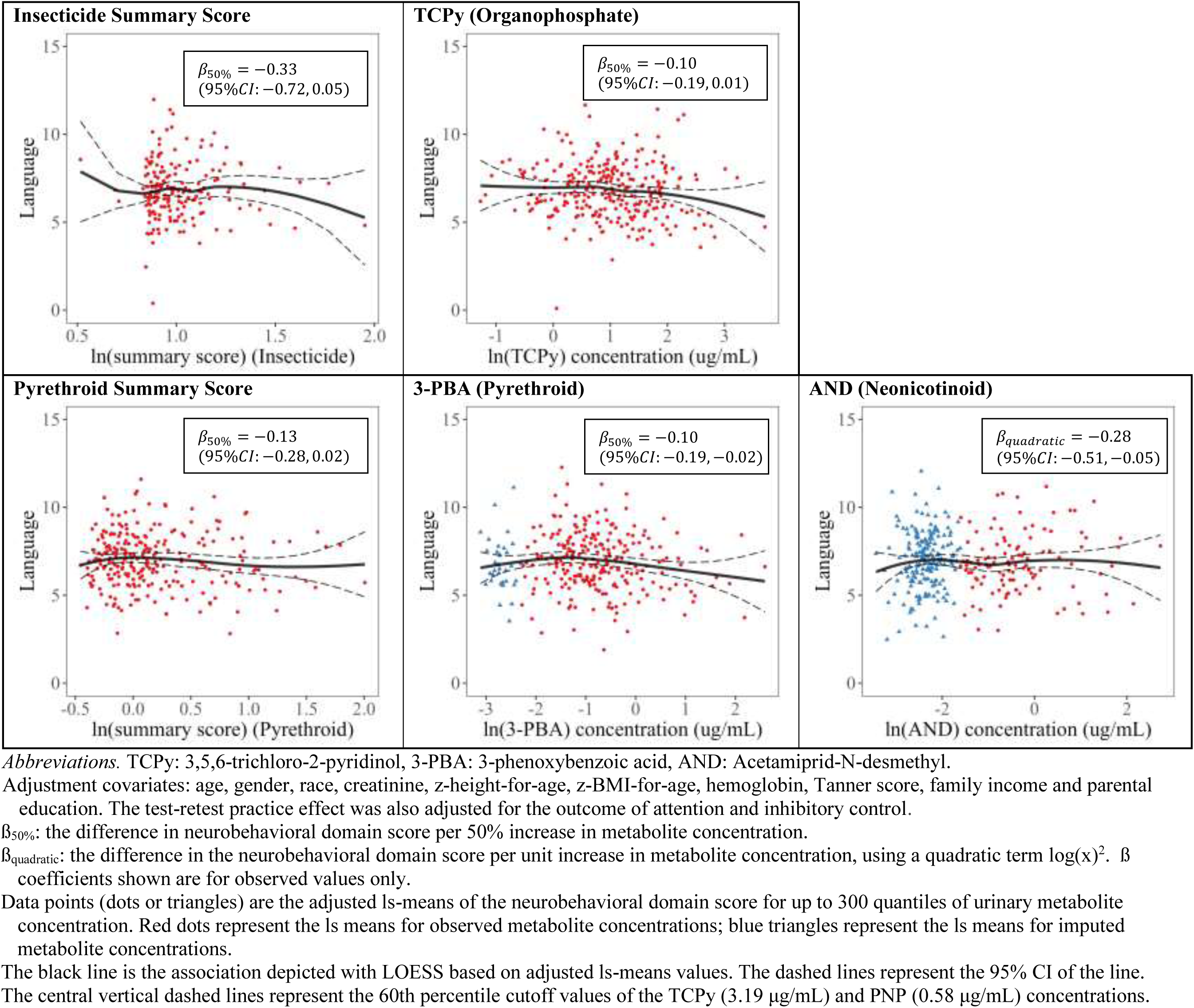
Language domain associations with log-transformed insecticide biomarkers; select associations with p values < 0.1.

**Figure 3.**
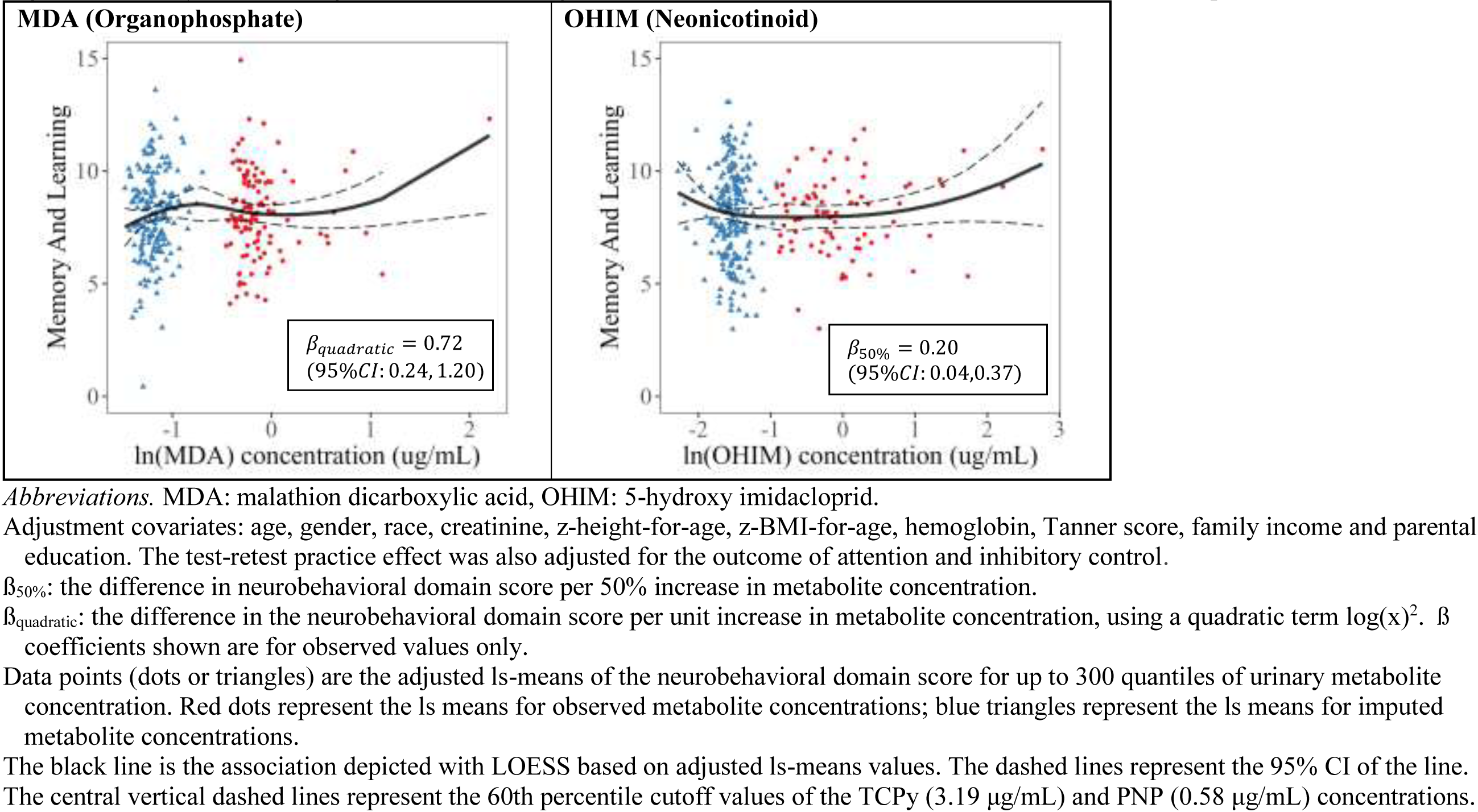
Memory and Learning associations with log-transformed insecticide biomarkers: select associations with p values < 0.1.

**Figure 4.**
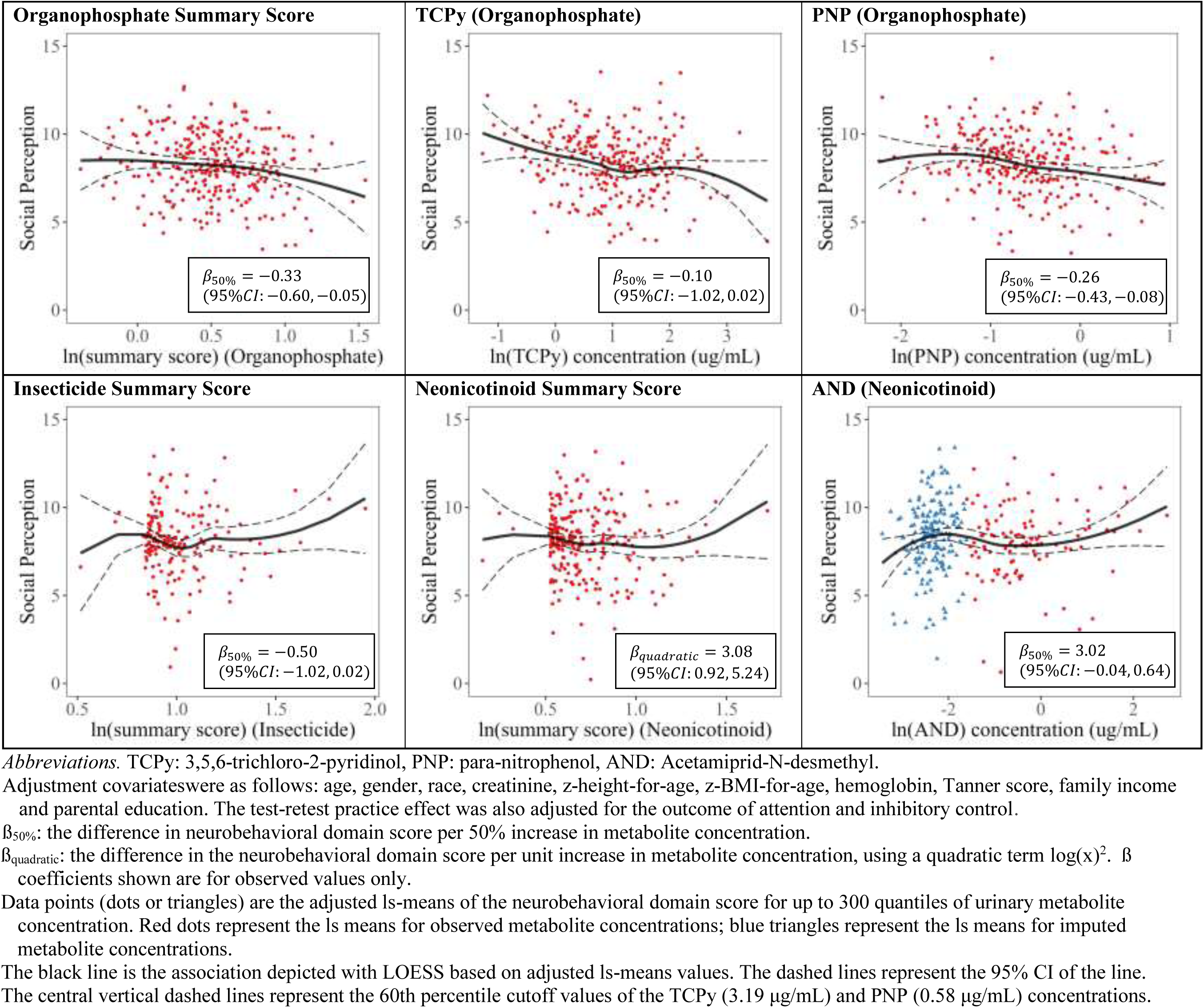
Social Perception associations with log-transformed insecticide biomarkers: select associations with p values < 0.1.

**Table 3.** Associations between urinary insecticide metabolite concentrations and neurobehavioral domain performance.

| | Difference in neurobehavioral domain score per 50% increase in metabolite concentration, $\beta_{50\%}$ (95% CI) | | | | |
| --- | --- | --- | --- | --- | --- |
|  | Attention and Inhibitory Control | Language | Memory and Learning | Visuospatial Processing | Social Perception |
| **Overall Insecticide Summary Score (n=520)** | -0.13 (-0.56, 0.30) | -0.33 (-0.72, 0.05)** | 0.28 (-0.17, 0.73) | 0.31 (-0.15, 0.78) | -0.50 (-1.02, 0.02)** |
| **Organophosphate Summary Score (n=520)** | -0.10 (-0.36, 0.17) | -0.11 (-0.36, 0.14) | 0.18 (-0.12, 0.48) | 0.12 (-0.19, 0.43) | -0.33 (-0.60, -0.05)* |
| TCPy Observed (n=520) | -0.02 (-0.12, 0.08)<sup>a</sup> | -0.10 (-0.19, -0.01)* | 0.05 (-0.06, 0.16) | 0.02 (-0.10, 0.13) | -0.10 (-0.22, 0.01)** |
| PNP Observed (n=515) | -0.12 (-0.26, 0.02)** | -0.08 (-0.23, 0.07) | 0.01 (-0.16, 0.17) | -0.13 (-0.31, 0.05) | -0.26 (-0.43, -0.08)* |
| MDA Observed (n=196) | -0.06 (-0.44, 0.33) | -0.00 (-0.30, 0.30) | 0.15 (-0.29, 0.58)<sup>b</sup> | 0.09 (-0.24, 0.42) | -0.18 (-0.50, 0.13) |
| MDA Imputed (n=324) | -0.04 (-0.28, 0.20) | -0.02 (-0.26, 0.23) | 0.06 (-0.22, 0.33) | 0.05 (-0.22, 0.32) | -0.08 (-0.38, 0.21) |
| **Pyrethroid Summary Score (n=520)** | -0.09 (-0.26, 0.08) | -0.13 (-0.28, 0.02)** | -0.02 (-0.19, 0.15) | 0.05 (-0.12, 0.22) | -0.11 (-0.32, 0.11) |
| 3-PBA Observed (n=438) | -0.04 (-0.13, 0.06)<sup>c</sup> | -0.10 (-0.19, -0.02)* | 0.00 (-0.10, 0.10) | 0.02 (-0.08, 0.11) | -0.05 (-0.17, 0.07) |
| 3-PBA Imputed (n=82) | -0.07 (-0.25, 0.1) | 0.04 (-0.14, 0.21) | -0.03 (-0.24, 0.18) | -0.04 (-0.27, 0.20) | -0.14 (-0.33, 0.06) |
| **Neonicotinoid Summary Score (n=520)** | 0.11 (-0.24, 0.46) | -0.12 (-0.45, 0.21) | 0.20 (-0.22, 0.62) | 0.07 (-0.30, 0.45) | -0.18 (-0.66, 0.30)<sup>d</sup> |
| AND Observed (n=191) | 0.14 (-0.00, 0.29)**<sup>e</sup> | 0.09 (-0.05, 0.22)<sup>f</sup> | 0.04 (-0.09, 0.17) | 0.02 (-0.11, 0.16) | 0.12 (-0.02, 0.26)** |
| AND Imputed (n= 329) | 0.01 (-0.08, 0.09) | 0.00 (-0.08, 0.09) | 0.01 (-0.09, 0.11) | 0.02 (-0.08, 0.13) | 0.02 (-0.09, 0.12) |
| OHIM Observed (n=137) | 0.03 (-0.13, 0.18) | -0.12 (-0.28, 0.05) | 0.20 (0.04, 0.37)* | -0.08 (-0.26, 0.10) | -0.01 (-0.28, 0.26) |
| OHIM Imputed (n=383) | -0.02 (-0.14, 0.10) | -0.01 (-0.12, 0.11) | -0.01 (-0.15, 0.13) | 0.01 (-0.13, 0.14) | 0.01 (-0.13, 0.14) |

Abbreviations. TCPy: 3,5,6-trichloro-2-pyridinol, PNP: para-nitrophenol, MDA: malathion dicarboxylic acid, 3-PBA: 3-phenoxybenzoic acid
AND: Acetamiprid-N-desmethyl, OHIM: 5-Hydroxy imidacloprid.
“Observed” are those with concentrations above the limit of detection (LOD). Imputed reflect estimated values using multiple imputation for concentrations below the LOD. Coefficients for both observed and imputed metabolites were derived from the same statistical model, with each term having its own distinct intercept.
Adjustments: age, gender, race, creatinine, z-height-for-age, z-BMI-for-age, hemoglobin, tanner score, salary, parental education. For Attention & Inhibitory Control, the model further adjusts for test-retest practice effect.
\*linear p p-value <0.05. \*\*linear p-value <0.1.
<sup>a</sup> Curvilinear model: TCPy $\beta$ : 0.39 (95% CI: -0.08, 0.85, p = 0.10), $\beta_{\text{quadratic}}$ : -0.21 (95% CI: -0.41, 0.00, p = 0.05).
<sup>b</sup> Curvilinear model: MDA $\beta$ : -0.56 (95% CI: -1.83, 0.71, p = 0.39), $\beta_{\text{quadratic}}$ : 0.72 (95% CI: 0.24, 1.20, p < 0.01).
<sup>c</sup> Curvilinear model: 3-PBA $\beta$ : -0.23 (95% CI: -0.48, 0.01, p = 0.07), $\beta_{\text{quadratic}}$ : -0.23 (95% CI: -0.37, -0.08, p < 0.01).
<sup>d</sup> Curvilinear model: Neonicotinoid summary score $\beta$ : -5.77 (95% CI: -9.92, -1.63, p < 0.01), $\beta_{\text{quadratic}}$ : 3.08 (95% CI: 0.92, 5.24, p < 0.01).
<sup>e</sup> Curvilinear model: AND $\beta$ : 0.44 (95% CI: 0.12, 0.77, p = 0.01), $\beta_{\text{quadratic}}$ : -0.35 (95% CI: -0.57, -0.13, p < 0.01).
<sup>f</sup> Curvilinear model: AND $\beta$ : 0.28 (95% CI: -0.05, 0.61, p = 0.09), $\beta_{\text{quadratic}}$ : -0.28 (95% CI: -0.51, -0.05, p = 0.02).

Among pyrethroids, the pyrethroid summary score and 3-PBA were inversely associated with Language (β_50%_ = -0.13 [95% CI: -0.28, 0.02, p=0.09 and β_50%_ = -0.10 [95% CI: -0.19, -0.02, p=0.02], respectively; Table 3). Additionally, 3-PBA had a significant negative curvilinear association with Attention & Inhibitory Control (ß_quadratic_ = -0.23 [95% CI: -0.37, -0.08, p<0.01], Figure 1).

For neonicotinoids, the neonicotinoid summary score had no linear associations with any neurobehavioral domain but had a positive curvilinear association with Social Perception (ß_quadratic_ = 3.08 [95% CI: 0.92, 5.24, p<0.01]; Table 3, Figure 4). OHIM had a positive linear association with Memory & Learning (β_50%_ = 0.20 [95% CI: 0.04, 0.37, p=0.02], Figure 3), whereas AND had a borderline positive association with Social Perception (β = 0.12 [95% CI: -0.02, 0.26, p=0.08]) and Attention & Inhibitory Control (β = 0.14 [95% CI: -0.00, 0.29, p=0.05]). There was evidence of curvilinearity in the associations of AND with Attention & Inhibitory Control (ß_quadratic_ = -0.35 [95% CI: -0.57, -0.13, p<0.01]), in which domain performance decreased the most in the highest AND concentrations (Figure 1). AND also had a significant curvilinear association with Language (ß_quadratic_ = -0.28 [95% CI: -0.51, -0.05, p=0.02]). The imputed metabolites alone were not linearly associated with any of the neurobehavioral domains.

### Thresholds and effect modification by gender

We observed evidence of a threshold effect for the organophosphates, TCPy and PNP, with Attention & Inhibitory Control. TCPy and PNP concentrations above the 60^th^ percentile (3.19 µg/mL and 0.58 µg/mL, respectively) were inversely associated with Attention & Inhibitory Control (β_50%_ = -0.22 µg/mL [95% CI: -0.43, -0.00, p=0.05], β_50%_ = -0.26 µg/mL [95% CI: -0.51, -0.01, p=0.04], respectively), whereas no significant associations were observed when this cut-off was not accounted for (Figure 1).

We observed effect modification by gender on the association between TCPy and Visuospatial Processing (p_interaction_=0.02; boys: ß_50%_ = 0.13 [95%CI: -0.05, 0.32, p=0.16], girls: ß_50%_ = -0.08 [95%CI: -0.21, 0.06, p=0.26]. We also observed borderline significant effect modification by gender in the OHIM association with Social Perception; however, we did not find any significant associations after gender stratification (p_interaction_=0.07; boys: ß_50%_= 0.17 [95% CI: -0.14, 0.49, p=0.28], girls: ß_50%_= -0.28 [95% CI: -0.64, 0.09, p=0.14], Figure S1).

### Pesticide mixtures and neurobehavioral performance

Based on the model fit analyses of the PLS components (Table 4), we included only the first composite variables generated by the PLS models, as additional composites did not meaningfully improve the adjusted R-squared value of the model compared with the first composite variable, and they were not statistically significant. The loadings of pesticide metabolite composite 1 are presented in the left panel of Figure 5 in bar graphs. The red bar graph indicates that the pesticide metabolite contributes in the same direction as the composite, and the blue bar graph indicates that it contributes in the opposite direction. The right panel in Figure 5 depicts the linear model between the first composite variable and each inflammatory biomarker.

**Figure 5.**
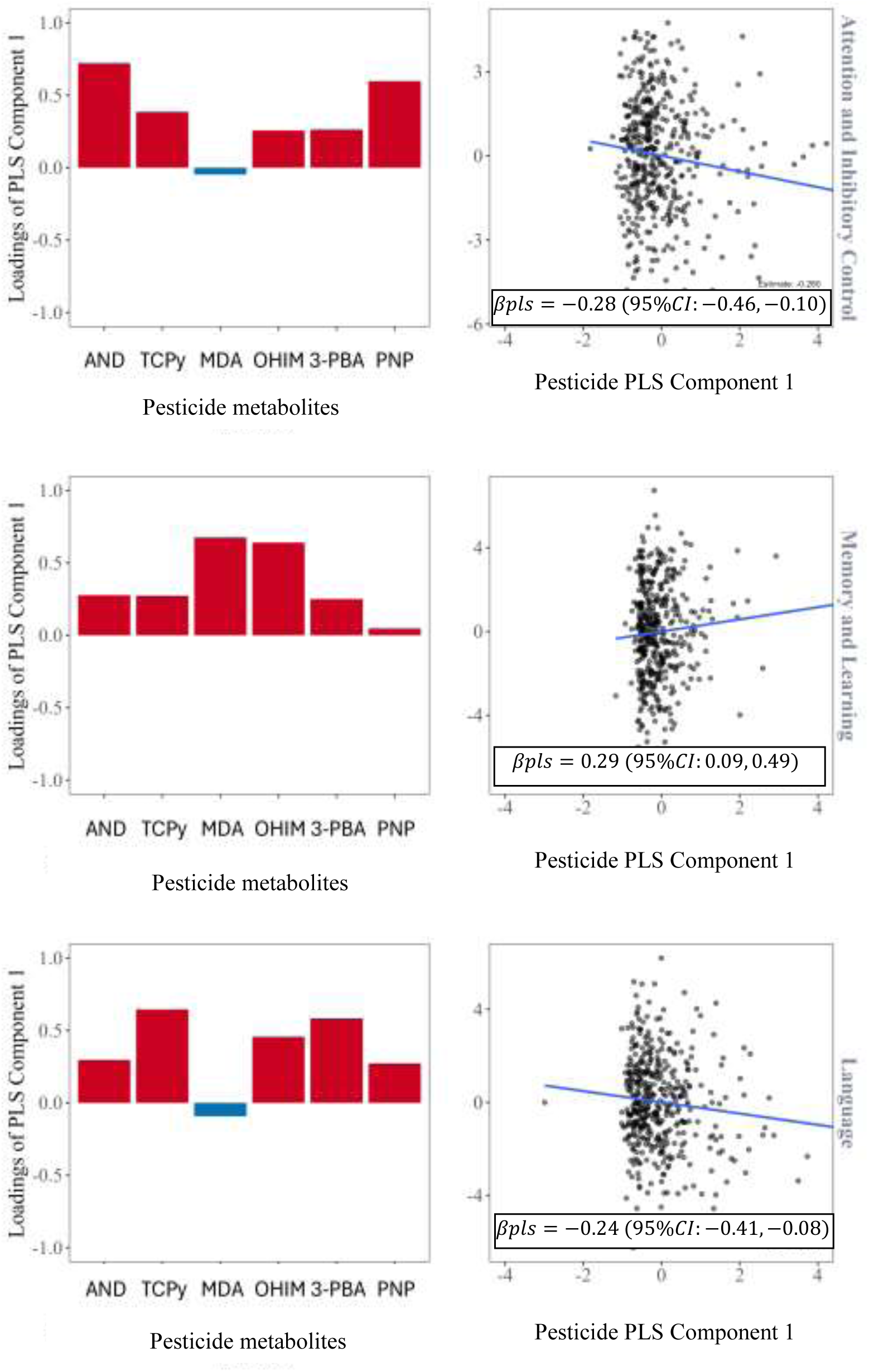
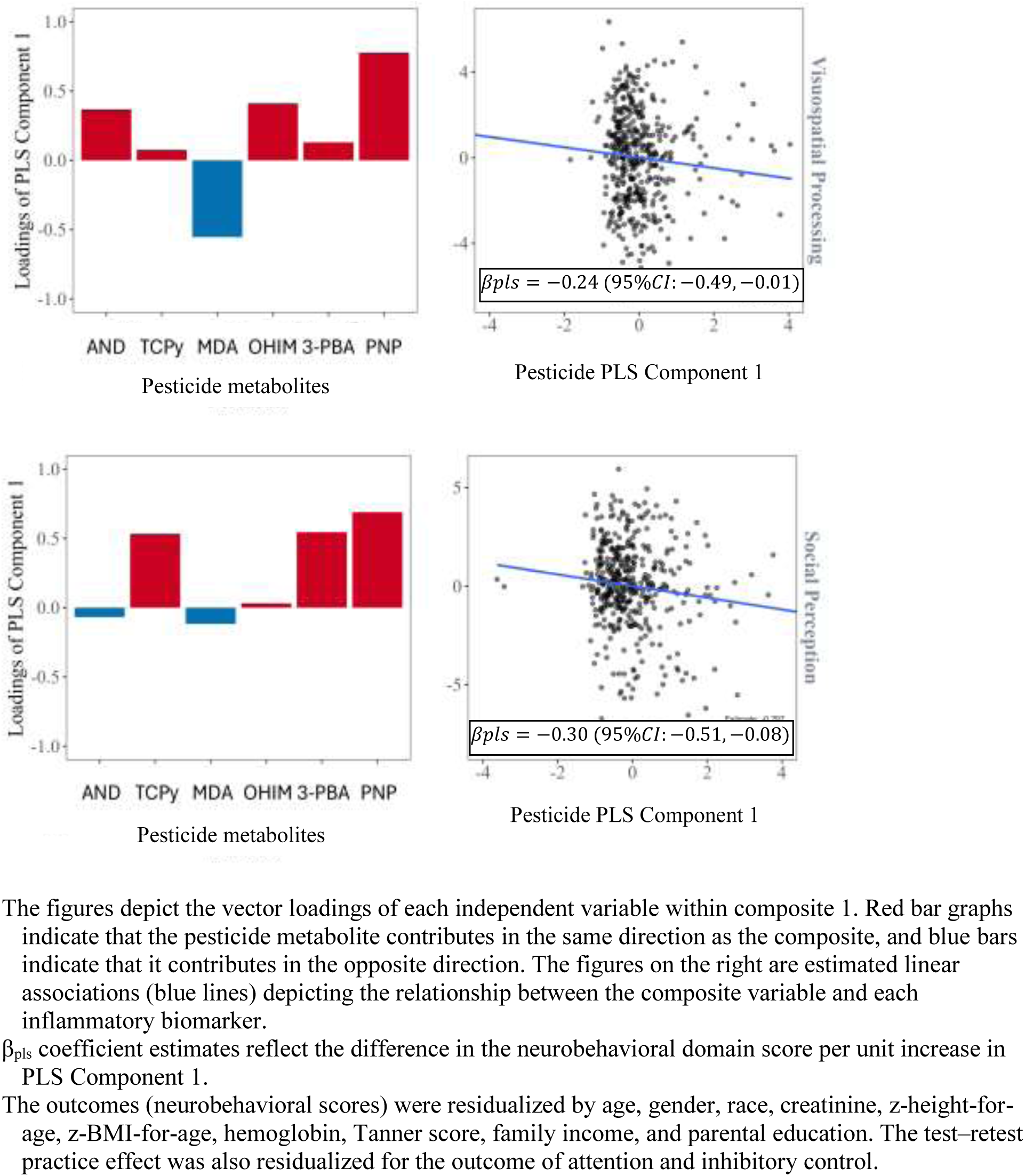
PLS component 1 loadings and associations between pesticide component 1 and neurobehavioral domains.

**Table 4.** Adjusted R^2^ values for the associations between urinary insecticide metabolite concentrations and neurobehavioral domain performance in adolescents (N=520)

| Neurobehavioral Domain | Full model | PLS |  |
| --- | --- | --- | --- |
|  |  | Component 1 | Component 1 + Component 2 |
| Attention and Inhibitory Control | 0.0047 | **0.0121** | 0.0129 |
| Language | 0.0101 | **0.0169** | 0.0184 |
| Memory and Learning | 0.007 | **0.0168** | 0.0161 |
| Visuospatial Processing | -0.0021 | **0.0072** | 0.0067 |
| Social Perception | 0.0054 | **0.0148** | 0.0139 |

The full model represents a linear regression model including all insecticide variables (TCPy, PNP, MDA, 3-PBA, AND, OHIM). The PLS models shown include the 1<sup>st</sup> component only and a combination of the 1<sup>st</sup> and 2<sup>nd</sup> PLS components. The outcomes are residualized for the following covariates: gender, age, race, creatinine, hemoglobin, z-height-for-age, z-BMI-for-age, Tanner score, family income and parental education. The bold text indicates statistical significance at the 95% confidence level.

Overall, the pesticide mixture was negatively associated with Attention & Inhibitory Control (difference in neurobehavioral domain score per unit increase in pesticide PLS Component 1 [β_pls1_]= -0.28, [95%CI: - 0.46, -0.10, p<0.01]), Language (β_pls1_= -0.24, [95%CI: -0.41, -0.08, p<0.01]), Visuospatial Processing (β_pls1_= -0.24, [95%CI: -0.48, -0.01, p=0.04]) and Social Perception (β_pls1_= -0.30, [95% CI: -0.51, -0.08, p<0.01]), whereas a positive association was observed with Memory & Learning (β_pls1_= 0.29, [95%CI: 0.09, 0.49, p<0.01]). Mixture modeling results were mostly consistent with the single pesticide metabolite analyses. The loadings of composite 1 variable showed that Attention & Inhibitory Control, Language and Visuospatial Processing were negatively associated with most of the urinary metabolites except for MDA. For Social Perception, the loadings indicated negative associations for TCPy, PNP and 3-PBA which is close to the result of models for single metabolites. Memory & Learning was positively associated with the conglomerate of metabolites.

### Mediation by inflammation biomarkers

Mediation by the inflammation marker summary score for the associations between neurobehavior outcomes and the 1st component generated by PLS was not statistically significant. The percentages of indirect effects over total effects were less than 0.01 (absolute value) for all tested associations (Table S1).

## Discussion

ESPINA is among the largest prospective cohort studies to assess the developmental effects of environmental pesticide exposure in adolescence. In this population, we previously reported that residential proximity to flower plantations (Friedman et al., 2020) and pesticide application periods (Espinosa da Silva et al., 2021; Suarez-Lopez et al., 2017a) are linked with greater pesticide exposure and lower neurobehavioral performance in research participants. In our current study, we delved deeper into the neurobehavioral associations of individual insecticide class exposures. Mixtures of all organophosphate, pyrethroid and neonicotinoid insecticides were associated with significantly lower performance in 4 out of 5 domains assessed, including: Attention & Inhibitory Control, Language, Social Perception and Visuospatial Processing. In contrast, the pesticide mixture was associated with greater performance in Memory & Learning. When analyzing pesticide metabolites individually, we found organophosphates to be inversely associated with Language and Social Perception, with evidence of a threshold effect for Attention & Inhibitory Control. We also identified quadratic relationships between organophosphates and Attention & Inhibitory Control as well as Memory & Learning. Pyrethroids demonstrated inverse associations with Language and quadratic associations with Attention & Inhibitory Control. Neonicotinoids had positive linear associations with Memory & Learning. Neonicotinoids also showed quadratic relationships with several neurobehavioral domains (Attention & Inhibitory Control, Language, Social Perception).

Our results regarding organophosphates and pyrethroids support our hypothesis that insecticides adversely impact neurobehavior in adolescents. However, the positive associations found for neonicotinoids were unexpected. In contrast to previous studies (Lee et al., 2020; Stein et al., 2016; Suarez-Lopez et al., 2017b, 2013; Wagner-Schuman et al., 2015; Yu et al., 2016), we did not find evidence that gender modifies the association between insecticides and Attention & Inhibitory Control. It is worth mentioning that higher insecticide exposure tertiles were associated with a greater percentage of mestizo compared to indigenous race, suggesting that mestizos experienced greater insecticide exposure than did indigenous populations. This may be partially explained by the fact that among our study participants, mestizo families lived closer to floricultural crops than indigenous families did (476 meters vs. 880 meters, respectively, p < 0.001). Further research is needed to understand how social and demographic factors impact secondary insecticide exposure.

The geometric mean concentrations of the organophosphates TCPy and MDA in our study were 3.3 and 1.5 times higher, respectively, than those of a nationally representative sample of 12–19-year-olds in the United States, as examined in the National Health and Nutrition Examination Survey (NHANES) (U.S. Department of Health and Human Services, 2020, 2019). In the NHANES, TCPy was measured from 2013-2014, and MDA was measured from 2009-2010. The geometric means of the neonicotinoids AND and OHIM were 1.72 and 1.71 times higher, respectively, than those of NHANES in 2015--2016 (U.S. Department of Health and Human Services, 2021). Given that Pedro Moncayo County is an agricultural community, it is reasonable to expect higher insecticide exposure levels among our study participants than among the general U.S. population. However, the geometric mean of PNP in the ESPINA population was quite similar (1.02 times greater) to the 2013--2014 NHANES concentrations (U.S. Department of Health and Human Services, 2020) and was only 1.37 times greater than the 2009--2010 NHANES geometric mean (U.S. Department of Health and Human Services, 2019). These results may have been confounded by the fact that PNP is not specific to organophosphates; it can also reflect exposure to other substances (e.g., acetaminophen and other drug precursors, fungicides, nitrobenzene, and certain dyes) (National Center for Biotechnology Information, 2021; Wessels et al., 2003). Interestingly, the geometric mean of 3-PBA was 17% lower among ESPINA participants than among NHANES 2013--2014 participants (U.S. Department of Health and Human Services, 2020).

### Organophosphates and neurobehavior

We observed a negative relationship of the organophosphate summary score with Social Perception and TCPy (a metabolite of chlorpyrifos and chlorpyrifos-methyl) had negative associations with Attention & Inhibitory Control (above the 60^th^ percentile threshold, 3.19 μg/mL), Language, and Social Perception (with borderline significance). These associations may be explained by alterations in AChE, serotonergic and dopaminergic systems. Like other organophosphates, chlorpyrifos and chlorpyrifos-methyl inhibit AChE in the central nervous system (Johnson et al., 2009; Kropp and Richardson, 2003; Rohlman et al., 2019; Skomal et al., 2021), which has been associated with impairments in language, memory, attention, inhibitory control and depression scores (Ismail et al., 2017b; Suarez-Lopez et al., 2013, 2019b, 2020). Chlorpyrifos also modifies serotoninergic (Aldridge et al., 2005a; Slotkin and Seidler, 2007) and dopaminergic (Aldridge et al., 2005b; Dam et al., 1999) neurologic activity, which may impact cognition and social behaviors (Tidey and Miczek, 1996; C. J. Yang et al., 2014) and increase the risk of ASD (Abdulamir et al., 2018; Chevallier et al., 2012; Chugani et al., 1999; Ernst et al., 1997; Gabriele et al., 2014; Scott-Van Zeeland et al., 2010; Staal, 2015). Developmental chlorpyrifos exposure has been linked to impaired mental development (Fluegge et al., 2016; Rauh et al., 2006), reduced social functioning (Guo et al., 2019), and ASD (Rauh et al., 2006; Von Ehrenstein et al., 2019). Similar to our findings, a study of adolescent insecticide applicators in Egypt revealed that higher urinary TCPy levels were associated with decreased attention, inhibitory control, and verbal functioning (Ismail et al., 2017b; Rohlman et al., 2014, 2019). Consistent with our finding of a threshold effect, a study in Mexico City identified an association between prenatal TCPy exposure and attention problems only at higher tertiles of exposure, with maternal urinary TCPy concentrations being similar to the urinary concentrations detected in our study (Fortenberry et al., 2014). PNP, a metabolite of parathion and methyl parathion, had consistent associations with TCPy. Like TCPy, PNP exhibited a negative association with Social Perception and had a negative association with Attention & Inhibitory control among participants with concentrations above the 60th percentile (0.58 μg/mL). Unlike TCPy, PNP was not associated with language performance. Although PNP’s behavioral effects in humans are not well studied (Garcia et al., 2003), children with detectable amounts of urinary PNP have been found to perform worse in terms of memory, attention, and behavior than unexposed children do (Ruckart et al., 2004). In animals, PNP- and parathion-containing compounds cause behavioral impairments (Edwards and Tchounwou, 2005; Gupta et al., 1985; Johnson et al., 2009), including autism-like behaviors (Song et al., 2021). Our findings regarding PNP’s threshold effect are consistent with those of rodent studies in which memory deficits (Johnson et al., 2009) and neurologic symptoms (Schulz et al., 1990; Shailesh Kumar and Desiraju, 1992) occurred only above certain exposure levels (oral administration of 0.2–0.9 mg/kg daily) (Johnson et al., 2009; Schulz et al., 1990; Shailesh Kumar and Desiraju, 1992). Given the negative impacts of both TCPy and PNP on Social Perception, it is unsurprising that the organophosphate summary score also correlated negatively with Social Perception performance. MDA, a metabolite of malathion, showed a weak positive quadratic association with Memory and Learning. Although the significance of this finding is unclear, particularly because only 38% of participants had detectable levels, it may complement previous studies on malathion’s neurobehavioral effects. Like other organophosphates, malathion inhibits neurologic AChE activity (del-Rahman et al., 2004; Fortunato et al., 2006; Kwong, 2002), but compared with other organophosphates, malathion is considered less toxic and has distinct pharmacokinetics (Agency for Toxic Substances and Disease Registry, 2003; Hoffmann and Papendorf, 2006; Nolan et al., 1984). In our study population, the urinary MDA concentration showed some evidence of AChE inhibition, although the results were not statistically significant (Skomal et al., 2021). In rodents, malathion was found to impair learning and memory (dos Santos et al., 2016; Geraldi et al., 2008; Lumsden et al., 2020; Valvassori et al., 2007) even at low levels, which did not affect neural AChE activity (dos Santos et al., 2016; Lumsden et al., 2020; Valvassori et al., 2007).

### Pyrethroids and neurobehavior

The pyrethroid summary score and 3-PBA were inversely associated with Language, and 3-PBA also had a negative curvilinear association with Attention & Inhibitory Control. As a nonspecific pyrethroid metabolite, 3-PBA can reflect exposures to Permethrin, Cypermethrin, Deltamethrin, Allethrin, Resmethrin, and Fenvalerate (Barr et al., 2010). 3-PBA’s alteration of dopaminergic activity (Carloni et al., 2012; Elwan et al., 2006; Nasuti et al., 2013, 2007; Richardson et al., 2015) has been strongly linked to ADHD symptoms (Cheon et al., 2005; Dougherty et al., 1999; Krause et al., 2006, 2000; Richardson et al., 2015) and hindered language acquisition (Wong et al., 2012). Prenatal exposure to 3-PBA and its precursors (Dalsager et al., 2019; Eskenazi et al., 2018; Fluegge et al., 2016; Furlong et al., 2017) has been associated with increased rates of ADHD (Dalsager et al., 2019), behavioral dysfunction (Furlong et al., 2017), and stunted mental (Fluegge et al., 2016) and language development (Eskenazi et al., 2018). Among children aged 2--9 years, urinary 3-PBA has been linked to behavioral and hyperactivity issues (Oulhote and Bouchard, 2013; Viel et al., 2017; Wagner-Schuman et al., 2015), ADHD (Lee et al., 2020; Richardson et al., 2015; Wagner-Schuman et al., 2015), reduced processing speeds (van Wendel de Joode et al., 2016), and diminished memory and verbal comprehension (Wang et al., 2016). Our findings support the findings observed in these studies.

### Neonicotinoids and neurobehavior

Our study revealed that neonicotinoid metabolites are positively associated with several neurobehavioral domains. The neonicotinoid summary score had a positive quadratic association with Social Perception, OHIM (from imidacloprid) was positively associated with Memory & Learning, and AND (from acetamiprid) had borderline positive associations with both Attention & Inhibitory Control and Social Perception. These positive associations may be explained in part by similarities between neonicotinoid insecticides and nicotine. Both substances excite nAChRs (Bal et al., 2010; Buszewski et al., 2019; Houchat et al., 2020; Kimura-Kuroda et al., 2012), a mechanism that is thought to mediate the short-term enhancing effects of nicotine on memory and attention (Campos et al., 2016; Jarvik, 1991; Nieradko-Iwanicka et al., 2019; Valentine and Sofuoglu, 2017; Wolter et al., 2019). Clinical trials have shown that nicotine administration to adult nonsmokers or smokers results in better performance in tests evaluating fine motor ability, attention and response time, and both short-term episodic and working memory (Heishman et al., 2010). Notably, however, at the highest concentrations of AND, the performance in Attention & Inhibitory Control was lower, as reflected by the negative quadratic term and visualized in Figure 1. Therefore, other non-nicotinic receptor-related effects associated with lower performance may be at play. The available data on AND and acetamiprid suggest that they have neurotoxic effects (Aagaard et al., 2013; Kagawa and Nagao, 2018; Sano et al., 2016; Shamsi et al., 2021; Thompson et al., 2020). In children and adults, urinary acetamiprid metabolites have been associated with neurologic symptoms such as memory loss, finger tremor, and headache (Marfo et al., 2015). Research assessing the mental health effects of neonicotinoid exposure in humans is scarce; hence, the neurobehavioral effects of neonicotinoid insecticides in both pediatric and adult populations are poorly understood. While the neurobehavioral effects of postnatal neonicotinoid exposure in humans remain unclear (Abreu-Villaça and Levin, 2017; Thompson et al., 2020), prenatal exposure has been linked to ASD (Keil et al., 2014), lower IQ (Gunier et al., 2017), and congenital defects such as anencephaly (W. Yang et al., 2014). Our study is among the first to characterize associations with neurobehavior via specific urinary biomarkers of neonicotinoid exposure. Further research in various agricultural populations is warranted.

Our exploration of peripheral blood inflammatory markers as mediators of insecticide–neurobehavior associations did not reveal any significant signals. However, considering the complexity of insecticide exposure and the possibility that different neurobehavioral domains may be impacted in varying ways, the role of inflammation as a biological mechanism warrants more thorough investigation. Inflammation may influence specific neurobehavioral outcomes differently depending on the type and level of insecticide exposure and is likely at different stages of life. The effects of inflammation on neurobehavioral outcomes can be modulated by various factors, including age-related changes in immune function, which are known to vary significantly from childhood through adulthood and into old age (Piber et al., 2019; Saavedra et al., 2023). The relationship between inflammation and neurobehavioral outcomes might be particularly complex in younger populations, where the developing brain is more vulnerable to environmental stressors, than in older adults, where chronic inflammation becomes more prevalent and could exacerbate neurodegenerative processes related to exposure. Research has shown that inflammatory markers often increase with age, particularly in older adults, where low-grade chronic inflammation becomes more pronounced. Markers such as IL-6, C-reactive protein (CRP), and TNF-α are commonly elevated in older populations and are associated with various age-related diseases, such as cardiovascular disease, diabetes, and cognitive decline (Piber et al., 2019; Saavedra et al., 2023). To gain a deeper understanding of these potential biological pathways, it is essential to conduct systematic and longitudinal studies. Such research would allow us to capture the dynamic nature of inflammatory responses and their relationship with neurobehavioral performance over time, across various domains, and in response to fluctuating exposure levels.

Our study has several strengths. This is one of the largest studies in adolescents to characterize the associations between pesticide exposure and neurobehavioral performance. Additionally, this is a population-based cohort study of participants in agricultural settings, which represent vulnerable populations that are exposed to many agrochemicals and are often not included in research. Additional strengths of the study include the comprehensive neurobehavioral performance assessments employed and the use of specific urinary biomarkers for organophosphate, pyrethroid, and neonicotinoid insecticides. An additional strength is that both the associations of individual biomarkers and mixtures of pesticide biomarkers with neurobehavioral outcomes were characterized. As agricultural crops are frequently sprayed with various chemicals or mixtures of chemicals, these analyses may more closely reflect the relationship between background exposure to pesticides and neurobehavioral performance.

Limitations of our study include the cross-sectional design of the present exposure and outcome constructs. Additionally, spot measurements of urinary insecticide metabolite concentrations may not adequately reflect chronic exposure. Hence, the negative associations of pesticide exposure with neurobehavioral performance presented in our analyses may reflect concurrent alterations in neurobehavioral performance. Urinary insecticide concentrations are known to vary across seasons (Attfield et al., 2014; Crane et al., 2013; Ismail et al., 2017b; Klimowska et al., 2020). In Pedro Moncayo County, periods of peak pesticide use coincide with increased floricultural production between October and May (Suarez-Lopez et al., 2017a). It is plausible that pesticide exposure from para-occupational sources may have been lower during the period in which we examined participants (July to October) than during the remainder of the year. Consequently, stronger associations between pesticide exposure and neurobehavioral outcomes might be observed during the peak floricultural production periods, as suggested by our previous work in this population (Espinosa da Silva et al., 2021; Suarez-Lopez et al., 2017a). This topic warrants further research. Although our current results align with studies linking stable exposure indicators (such as residential proximity to flower plantations (Friedman et al., 2020) and AChE activity (Suarez-Lopez et al., 2013)) to neurobehavioral deficits, a more comprehensive understanding of the long-term consequences could be achieved by measuring insecticide concentrations and neurobehavioral performance at multiple time points throughout the year. Finally, although measuring urinary biomarkers of pesticide exposure allows us to characterize specific exposures, such measures are not always stable considering the short half-lives of many modern pesticides. The urinary concentrations of organophosphates, pyrethroids, and neonicotinoids can differ with season and insecticide application period (Attfield et al., 2014; Crane et al., 2013; Han et al., 2018; Ismail et al., 2017b, 2017a; Klimowska et al., 2020; Saillenfait et al., 2015), day to day, or even within one day (Attfield et al., 2014; Bradman et al., 2013; Egeghy et al., 2011; Griffith et al., 2011; Li et al., 2019; Wolfe et al., 1970). While pyrethroid metabolites such as 3-PBA tend to be more stable over time than organophosphates are (Egeghy et al., 2011; Klimowska et al., 2020; Wielgomas, 2013), conflicting data exist (Attfield et al., 2014; Morgan et al., 2016). Among neonicotinoids, spot urinary samples of AND were moderately reliable for estimating exposure over 44 days (Li et al., 2020). In comparison, the urinary concentrations of imidacloprid and its metabolites had poor reproducibility during 44-day (Li et al., 2020) and one-year periods (Klimowska et al., 2020). Multiple urine samples over time are often needed to account for individual variability and to better reflect long-term exposure (Attfield et al., 2014; Bradman et al., 2013; Griffith et al., 2011; Li et al., 2020; Wessels et al., 2003), but this strategy comes at a substantial financial and logistical cost.

Further research is needed to identify reliable biomarkers of exposure to different insecticide classes, optimal measurement methods, and the applicability of these biomarkers to acute versus chronic exposure.

## Conclusion

Our research provides insights into the effects of exposure to insecticides (organophosphates, pyrethroids, and neonicotinoids) on the neurobehavioral performance of adolescents in an agricultural region of Ecuador. Our results show that background exposure to organophosphates, pyrethroids, and neonicotinoids can impact different aspects of neurobehavioral performance, reinforcing evidence that each insecticide subtype within these classes has distinct biological effects (Matsuda et al., 2009; Roberts and Karr, 2012; Slotkin et al., 2008). We found consistent associations of organophosphate urinary metabolites with neurobehavioral alterations, which concurs with past research. We observed that organophosphate biomarkers had negative linear associations with the domains of Language (TCPy), Social Perception (organophosphate summary score, PNP and TCPy), and Attention & Inhibitory Control (TCPy and PNP, but only above the 60^th^ percentile threshold). Although there was evidence of effect modification by gender in the TCPy-Visuospatial Processing association, we did not find significant associations after gender stratification. The pyrethroid metabolite 3-PBA had a negative linear association with Language and a negative curvilinear association with Attention & Inhibitory Control. These negative associations of organophosphate and pyrethroid exposure with neurobehavioral performance are consistent with previous population-based studies, most notably with attention and inhibitory control and language alterations. Contrasting these findings, neonicotinoid insecticides were associated with better scores in Memory & Learning and Social Perception, which concurs with experimental evidence that the administration of nicotine, a substance that has physiological properties similar to those of neonicotinoid insecticides, can improve neurobehavioral performance in humans. In this cohort, we previously reported that pesticide exposure was positively associated with inflammatory markers and that such markers were negatively associated with neurobehavioral performance. However, our exploration of mediation by peripheral blood inflammatory markers in pesticide–neurobehavior associations did not reveal any notable signals in this group of adolescents. Since various inflammation markers often increase with age, it is advisable to replicate these findings in older populations, including age groups where low-grade chronic inflammation becomes more pronounced. Overall, our findings suggest that background insecticide exposure in agricultural settings is associated with alterations in neurobehavior in adolescents. We highlight the importance of limiting environmental insecticide exposure during developmental years and the need for further research on insecticide exposure and neurobehavior assessed multiple times within a year and across multiple seasons.

## Data Availability

All data produced in the present study are available upon reasonable request to the authors

https://knit.ucsd.edu/espina/

## Acknowledgements

Research reported in this publication was supported by the National Institute of Environmental Health Sciences (NIEHS) of the National Institutes of Health under Award Numbers R01ES030378, R01ES025792, R21ES026084, and U2CES026560 and the National Institute for Occupational Safety and Health of the National Institutes of Health under Award Number 1R36OH009402. Jessica Yen was funded by the Summer Research Fellowship at the UCSD School of Medicine. Funding was also provided by NIEHS grant K01ES031697 to Dr. Georgia Kayser. We thank ESPINA study staff, Fundación Cimas del Ecuador, the Parish Governments of Pedro Moncayo County, community members of Pedro Moncayo, and the Education District of Pichincha-Cayambe-Pedro Moncayo counties for their contributions to and support this project. We thank Dr. Rajendra Parajuli, for his contributions on this manuscript.

## Author contributions

**Jessica Yen**: Conceptualization, and original draft preparation **Kun Yang**.: Software, Data Analysis, and Visualization. **Xin M. Tu**: Validation, Reviewing and Editing. **Georgia Kayser**: Reviewing and Editing. **Ana Skomal**: Reviewing and Editing. **Sheila Gahagan:** Reviewing and Editing. **Jose Suarez-Torres**: data collection, reviewing and editing. **Suzi Hong**: Reviewing and Editing. **Raeanne C. Moore**: Reviewing and Editing. **Jose R. Suarez-Lopez**: Funding, conceptualization, data curation, data collection, original draft preparation.

---

**Table S1.** Coefficients of the indirect effect (mediation) of the inflammation summary score on the associations between the neurobehavioral domain and the corresponding pesticide metabolite PLS component.

| **Neurobehavioral Domain** | **Indirect Effect (95% CI), p value** | **Indirect/Total Effect** |
| --- | --- | --- |
| Attention and Inhibitory Control | 0.001 ([-0.006, 0.009], 0.71) | -0.005 |
| Language | -0.002 ([-0.013, 0.010], 0.76) | 0.006 |
| Memory and Learning | -6e-04 ([-0.006, 0.005], 0.82) | -0.002 |
| Visuospatial Processing | -4e-04 ([-0.006, 0.006], 0.90) | 0.001 |
| Social Perception | -0.002 ([-0.011, 0.008], 0.71) | 0.004 |

Adjustment covariates: age, gender, race, creatinine, z-height-for-age, z-BMI-for-age, hemoglobin, Tanner score, family income and parental education. The test-retest practice effect was also adjusted for the outcome of attention and inhibitory control.

**Figure S1.**
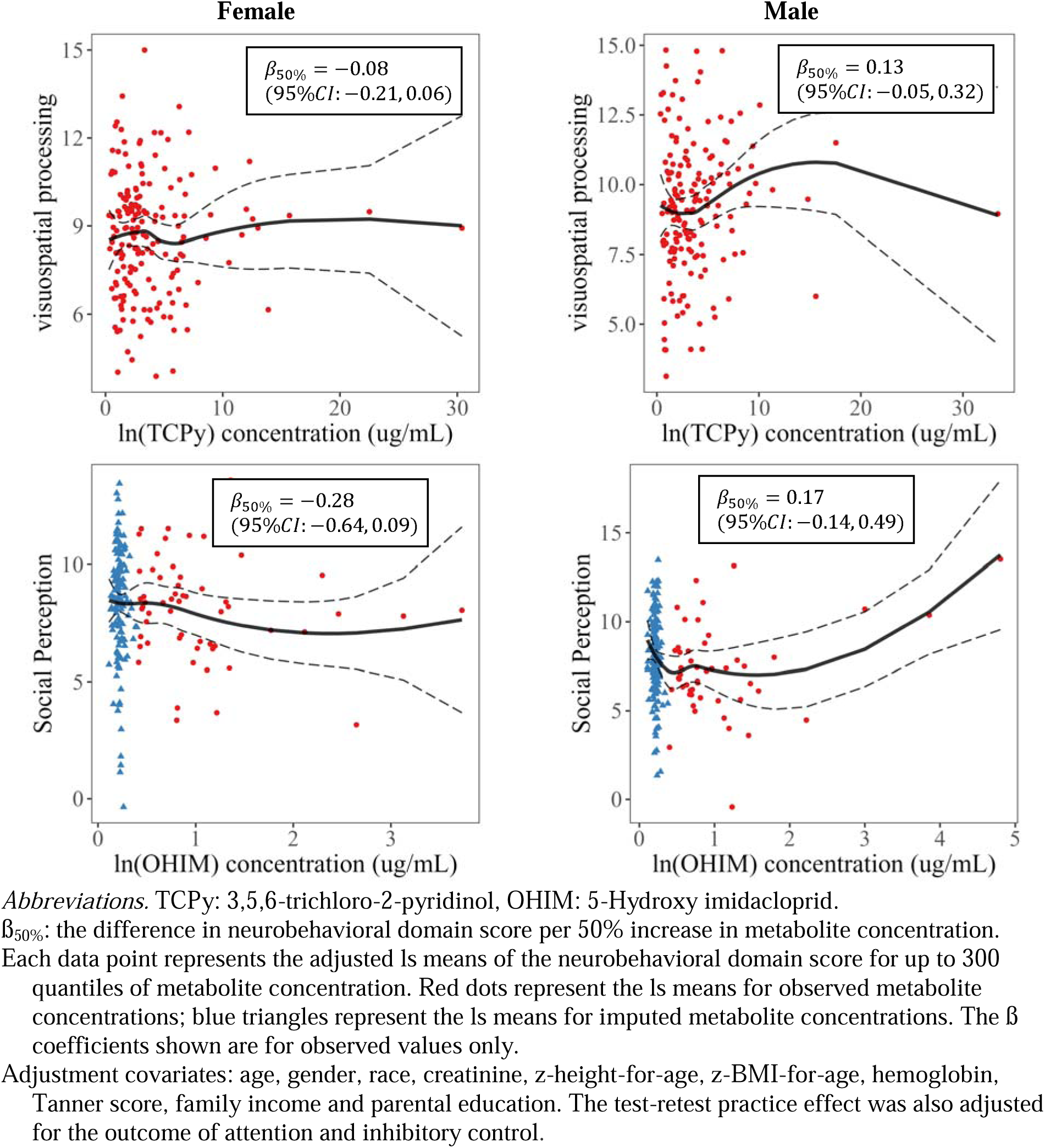
Gender-stratified associations of urinary insecticide metabolites with the domains of visuospatial processing and social perception.

